# Pickleball injuries, search interest, and participation in the United States, 2014-2023: an ecological time series study

**DOI:** 10.64898/2026.10.14.26365555

**Authors:** Daniel Seung Kim, Felipe Lobelo, Jared C. Durieux, Christos Colovos

## Abstract

**OBJECTIVE:** To describe (1) US emergency department-treated pickleball injury burden and the injury-to-participation measure among total players, with burden among adults aged 60 to 79 years as a descriptive subgroup, and (2) determine whether general internet search interest for “pickleball” is also associated with total pickleball-related injuries.

**DESIGN:** Ecological time series study, United States, 2014 to 2023.

**DATA SOURCES:** National Electronic Injury Surveillance System (NEISS) emergency department microdata yielded 1,738 adjudicated pickleball cases, 921 of them orthopedic, weighted to an estimated 101,450 injuries nationally. Google Trends, English Wikipedia pageviews, and GDELT news volume served as general pickleball attention signals, with the Trends index for “sourdough” as a negative control. Sports and Fitness Industry Association (SFIA) participation estimates supplied a total player denominator.

**MAIN OUTCOME MEASURES:** Annual national injury estimates with design-based 95 percent confidence intervals; injury-to-participation measures; and correlations between total pickleball-related injuries and general pickleball attention signals before and after removing trend and seasonality.

**RESULTS:** Estimated emergency department-treated pickleball injuries rose 17.5-fold, from 1,391 in 2014 (highly unstable) to 24,294 in 2023. 70 percent of adjudicated cases involved adults aged 60 to 79. During 2017-2023, injury estimates rose 4.0-fold and participation rose about 4.4-fold. No temporal trend was detected in the injury-to-participation measure (P=0.59). Search interest looked like an excellent proxy: in annual levels it correlated with total pickleball-related injuries at R=0.96, and Pickleball Wikipedia pageviews at R=0.98. However, so did searches for "sourdough", at R=0.76. Removing trend and seasonality left little (monthly residual R=0.084; year-on-year R=0.28 over 96 months, 2015-2023), and the seasons ran opposite, with injuries peaking in February and searches in July, such that, on 12-month mean profiles, the sourdough negative control fitted the injury season (R=0.96) while pickleball ran against it (R=-0.40). General pickleball search interest and pickleball news coverage remained correlated in year-on-year changes in 2017-2023 (R=0.64), whereas their corresponding correlations with total pickleball-related injuries in 2017-2023 were 0.25 and 0.20.

**CONCLUSIONS:** The absolute burden of emergency department-treated pickleball injury has grown substantially, but the available surveillance data cannot resolve whether the sport has become more hazardous per participant. The observed rise is consistent with growth in exposure alone. The strong correlation between search interest and injury counts was not specific, the association was substantially attenuated after detrending, and the seasonal patterns were opposed. These descriptive results do not establish that search interest can substitute for injury surveillance, and series that rise together should not be assumed to be related.

**WHAT THIS STUDY ADDS:**

- Injury estimates rose 4.0-fold and participation rose about 4.4-fold during 2017-2023. No temporal trend was detected using the total-player participation denominator. However, this does not establish unchanged risk per pickleball player.
- Strong annual correlations between total pickleball-related injuries and general internet attention for “pickleball” (R=0.96 for search interest, 0.98 for Wikipedia pageviews) were also observed for a sourdough negative control (R=0.76). Injury-search correlations were substantially attenuated after accounting for trend and seasonality.
- General pickleball search interest remained associated with pickleball news coverage in year-on-year changes in 2017-2023 (R=0.64); however, their year-on-year correlations with total pickleball-related injuries in 2017-2023 were smaller (R=0.25 and 0.20).
- Taken together, these analyses caution against interpreting rising pickleball-related injury counts as rising individual risk or using search interest alone to extrapolate to injury burden.

## INTRODUCTION

Pickleball participation and reported injuries have risen sharply in the United States (US) <u>[1]</u>. Analyses of the National Electronic Injury Surveillance System (NEISS) describe emergency department attendance climbing steeply, with fractures and sprains prominent, and much of the burden borne by adults aged 60 and over <u>[2–7]</u>. These counts establish the burden of injury, but they cannot alone distinguish more people playing from greater risk per player. That distinction requires a participation denominator <u>[8,9]</u>.

A separate question is whether internet attention can help measure the injury burden. Search interest is readily available, and a previous study reported an association between pickleball search volume and injury counts <u>[10]</u>. Yet searches may reflect publicity, in addition to participation or injury. Hence, two series that both rise over time can correlate even when neither causes the other (e.g., chocolate consumption and the number of Nobel laureates) <u>[11,12]</u>.

We therefore asked two questions: whether estimated injury rates changed over time when normalized to the total number of pickleball players, and whether general search interest for “pickleball” tracked total pickleball-related injuries after adjusting for trend and seasonality. We used the total-player participation denominator and compared search interest with Wikipedia pageviews, news coverage, and a negative control: sourdough, another growing enthusiasm but with no obvious route to a broken wrist <u>[13]</u>.

## METHODS

### Ethics statement

This study used publicly available, de-identified data. An external review by Bluebonnet Ethical Review determined that this activity is classified as Not Human Subjects Research (NHSR) and that IRB review is not required. Reporting follows STROBE (see **Supplementary Table S1**).

### Injury data and case definition

We conducted an ecological time series study using US data from 2014 to 2023. We obtained annual NEISS case-level files from the US Consumer Product Safety Commission (CPSC) <u>[14]</u>. NEISS draws a stratified probability sample from about 100 hospital emergency departments across five strata; case weights permit estimation of national injury totals <u>[15]</u>. Because pickleball has no dedicated NEISS product code, cases were identified from free-text narratives <u>[16]</u>.

A naive search for the string “pickle” returned 1,958 records. We reviewed these narratives and excluded 11 percent under the rule below, most of them injuries from pickle jars and food preparation (e.g., “injured wrist opening pickle jar”; “cut finger slicing a pickle”) rather than from the sport. We therefore required evidence of the game, either with an explicit “pickleball” (with common misspellings) or “pickle” alongside a playing term such as “court”, “paddle”, or “serve”, and we excluded food and jar narratives. The rule retained 1,739 of the 1,958 records (yield 88.8%). This yield is not a validated positive predictive value, because no independent gold-standard adjudication was performed. One retained record, a food-related kitchen injury in a three-year-old (case 140424871), was removed on adjudication, leaving 1,738 cases for analysis. All results below use this adjudicated set (see **Supplementary Table S2**).

We used all 1,738 adjudicated cases (total pickleball-related injuries) for injury counts, injury-to-participation analyses, and the main injury correlations, seasonality, and negative-control analyses. Subtype correlations are labelled by subtype where they are reported. Total pickleball-related injuries measure the burden and form the series a general “pickleball” search-interest proxy would have to track. The orthopedic subset (921 fractures, dislocations, and sprains/strains) is retained as a sensitivity check. Orthopedic injuries refer to fractures, dislocations, and sprains/strains by NEISS diagnosis code (**Supplementary Table S3**): the acute fall- and impact-related injuries expected to dominate emergency presentations in players aged 60 years or older. We converted NEISS age codes to years and mapped orthopedic diagnoses using NEISS diagnosis codes (fracture 57, dislocation 55, sprain/strain 64) per the CPSC NEISS Coding Manual (January 2023) (**Supplementary Table S4**).

### National estimates and their uncertainty

We calculated national estimates by summing case weights. Design-based variance using the NEISS stratum and primary sampling unit variables was computed using Taylor linearization on the full annual responding hospital frame, retaining hospitals with zero pickleball cases so that sampled primary sampling units are not dropped after domain subsetting. These public-data variances do not include CPSC response and calibration adjustments, and should be read as approximate design-based uncertainties, not official CPSC standard errors <u>[17]</u> used by CPSC and the CDC injury surveillance system (WISQARS) <u>[18]</u>. We computed a standard error for each estimate and reported the 95 percent confidence interval and coefficient of variation in the tables. We flagged each estimate against CPSC stability guidance, which specifies an estimated total of at least 1,200, at least 20 sample cases, and a coefficient of variation at or below 33 percent. Estimates missing any criterion are labeled ‘Unstable’ (**Table 1**); most annual estimates here fall in that category and should be read as order-of-magnitude signals.

**Table 1.** Adjudicated pickleball cases, NEISS 2014 to 2023, with design-based (Taylor linearization) 95 percent confidence intervals. Consumer Product Safety Commission (CPSC) stability guidance requires an estimated total of at least 1,200, at least 20 sample cases, and a coefficient of variation at or below 33 percent. Only the 2023 estimate meets all three thresholds. Annual estimates are rounded to whole injuries and sum to 101,451; the pooled total of 101,450 reported in the text was computed before rounding. Orthopedic-subset estimates (sensitivity series) are tabulated in **Supplementary Table S3**.

| Year | Sample cases (N) | National estimate | 95% CI | CV (%) | CPSC stability |
| --- | --- | --- | --- | --- | --- |
| 2014 | 24 | 1,391 | 127 to 2,655 | 46.4 | Unstable |
| 2015 | 60 | 4,374 | 69 to 8,680 | 50.2 | Unstable |
| 2016 | 68 | 4,991 | 917 to 9,065 | 41.6 | Unstable |
| 2017 | 97 | 6,072 | 78 to 12,065 | 50.4 | Unstable |
| 2018 | 131 | 8,310 | 0 to 16,680 | 51.4 | Unstable |
| 2019 | 172 | 11,168 | 0 to 22,835 | 53.3 | Unstable |
| 2020 | 145 | 9,029 | 0 to 18,303 | 52.4 | Unstable |
| 2021 | 280 | 14,825 | 0 to 30,246 | 53.1 | Unstable |
| 2022 | 288 | 16,997 | 3,006 to 30,988 | 42.0 | Unstable |
| 2023 | 473 | 24,294 | 10,539 to 38,049 | 28.9 | Stable |

Orthopedic sensitivity-check subset of injuries by year 2014 to 2023: 15, 32, 40, 59, 77, 92, 92, 162, 140, 212

### Participation denominators

Throughout, “older adults” means adults aged 60 and over, following the NEISS pickleball literature <u>[2–7]</u>. For 2017-2023, we divided annual national injury estimates by Sports and Fitness Industry Association (SFIA) participation estimates using a single denominator: total players (SFIA ages 6 and older) <u>[1,8,9]</u>. The SFIA estimates as used here are national totals without age detail and cover 2017–2023, so denominator-adjusted analyses are restricted to those years while injury counts are reported for 2014–2023. Contextual series extending beyond 2023 (court totals) describe the sport’s growth but enter no rate or correlation analysis. SFIA’s annual report estimates US sports participation from a national survey.

### Attention signals and contextual measures

We retrieved the monthly Google Trends index for general search interest in “pickleball” (US, January 2015 to December 2023, 108 months) and monthly English Wikipedia pageviews for the Pickleball article (102 complete months, July 2015 to December 2023). We recorded the exact query and retrieval date, because the Trends index is a sampled, rescaled quantity that varies between retrievals <u>[19,20]</u>. Both retrieved files were checked against their retrieval receipts and are included in the replication package. As a negative control, we retrieved the Trends index for “sourdough”, a craze that rose over the same years and is not associated with pickleball injuries. We used monthly pickleball news volume from GDELT, a global news index, for 2017-2023 to examine whether search interest tracked media attention <u>[21]</u>. USA Pickleball court totals provided context on the sport’s growth (the 2021 total of 38,140 was attributed to Places2Play in the 2022 fact sheet; later totals are attributed to Pickleheads) <u>[22]</u>. These sparse supply measures mark a real and rising sport, but were not treated as measures of play or injury. Monthly injury estimates covered 2014–2023 (120 months); attention-signal retrievals covered January 2015–December 2023 for Trends (108 months), July 2015–December 2023 for Wikipedia (102 months), and 2017–2023 for GDELT news (84 months). Participation denominators are annual only.

### Analysis

For the injury-to-participation measure (annual injuries per 1,000 players), we tested for a trend over the calendar year using ordinary least squares regression with t-based inference. The denominator is treated as fixed, so rate confidence intervals (**Supplementary Table S5**) are lower bounds on uncertainty. For attention analyses, we calculated Pearson correlation coefficients in levels, tested series for stationarity using the augmented Dickey-Fuller test with a constant (5 percent critical value, -2.9) <u>[23]</u>, and examined correlations in year-on-year changes. Level correlations were computed at both annual and monthly aggregation. Correlations are reported as descriptive point estimates without confidence intervals or significance tests, apart from the labelled nominal Spearman test in the sensitivity analysis; inference in this study rests on the injury-to-participation regression. Year-on-year correlations use 12-month differences, which remove a fixed repeating seasonal component; we did not smooth these differenced series.

In a sensitivity analysis, we repeated every monthly analysis using unweighted total case counts (each record counted once) rather than NEISS sampling weights; orthopedic unweighted results are reported separately where they appear. We also compared changes in pickleball cases and all records in the downloaded consumer-product NEISS files in 2020 (the only year in which the national injury estimate fell year-on-year (**Table 1**)) to contextualize the pandemic decline. Monthly analyses were descriptive, not confirmatory.

We used seasonal-trend decomposition based on loess (STL) <u>[24]</u>, with a robust outer loop, to separate trend, seasonality, and remainder, then compared (i) residual correlations and (ii) Pearson correlations of the standardized seasonal components across signals.

## RESULTS

### Injury burden and participation

Estimated emergency department-treated pickleball injuries rose 17.5-fold, from 1,391 in 2014 (highly unstable; **Table 1**) to 24,294 in 2023 (**Figure 1A**; annualized trends in **Supplementary Figure S1**). Adults aged 60 to 79 accounted for 70 percent of adjudicated cases. Annual coefficients of variation averaged about 47 percent over 2014-2023 under the full-frame variance calculation. Only the 2023 estimate met the stated CPSC stability criteria; the 2014 coefficient of variation was 46.4 percent (**Table 1**).

**Figure 1.**
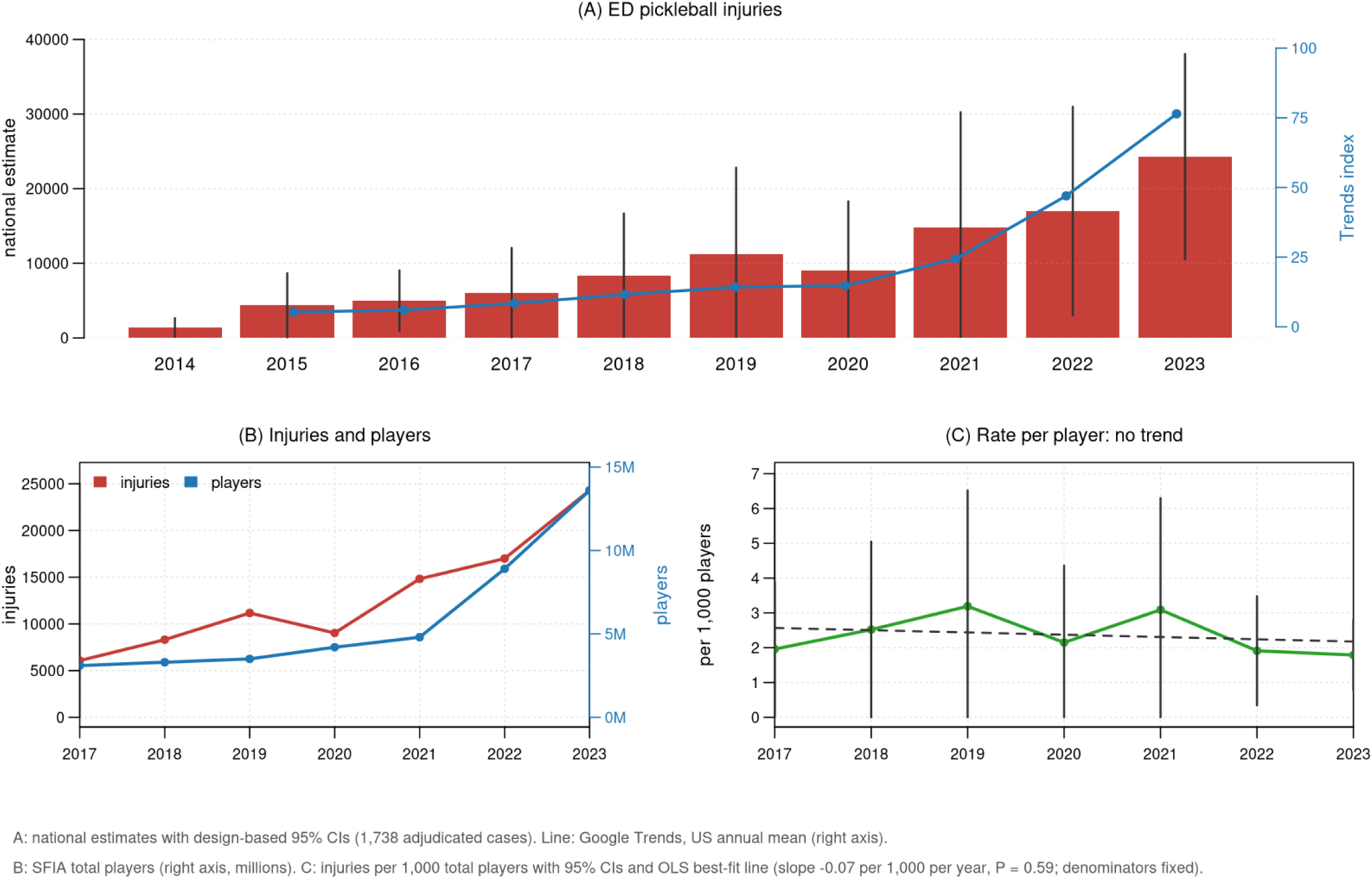
Pickleball injuries, players, and the injury-to-participation measure. (A) Annual national estimates of all pickleball injuries (1,738 adjudicated cases), with design-based 95 percent confidence intervals and Google Trends interest (right axis). Counts alone cannot separate rising participation from rising per-player risk; see panel C. Annualized levels with OLS best-fit trends are shown in **Supplementary Figure S1** (injury slope +2,154 cases/year; search +7.38 index points/year). (B) National injury estimates and SFIA total players, 2017 to 2023. (C) Injury-to-participation measure per 1,000 total players with 95% confidence intervals and OLS best-fit line. No temporal trend was detected (slope −0.07 per 1,000 per year, 95% CI −0.36 to 0.23; P=0.59; denominators treated as fixed).

During 2017-2023, national injury estimates rose 4.0-fold and participation rose about 4.4-fold (**Figure 1B**). Emergency departments treated more cases over time (P<0.001 over 2014-2023), a pattern consistent with more people playing, but no temporal trend was detected in the injury-to-participation measure (P=0.59; **Figure 1C**). Only the 2023 annual estimate met stability guidance. Repeating the injury-to-participation analysis on the orthopedic subset gave the same verdict (P=0.29; slope negative), with no upward drift in the orthopedic share of cases.

A total of 1,214 of 1,738 adjudicated cases (69.9%; 74.1% of weighted estimates) were in the 60–79 age group. Adults aged 80 years or older accounted for 98 cases (5.6%; 5.4% of weighted estimates). Among 472 fractures, code 34 (wrist) accounts for 30.5%, followed by code 79 (lower trunk; 15.7%) and code 33 (lower arm; 14.4%), with code 37 (ankle) at 3.2%. Fractures concentrate in ages 60–79 (367 of 472, 77.8%); body-part-specific rates by age are not computable here because participation denominators matched to the 60 to 79 age band are not available in the sources used.

Case-level splits support the main results. Adjudicated cases were evenly split by sex (875 male, 863 female; weighted 51,450 vs 50,000). The 60–79 band dominated every year (**Supplementary Figure S2**); cases under 60 rose fastest in proportional terms after 2020 (163 in 2023), while the 60–79 band had the largest absolute increase. Cases concentrated in 2019–2023 (1,358 of 1,738). By subtype, fracture and sprain series both show near-zero year-on-year search coupling (fracture R=−0.003, sprain R=0.062; levels 0.51 and 0.23, using all 108 calendar months and 96 year-on-year pairs with zero-case months retained). Thus, we detected no divergence suggesting a hidden risk signal in acute injuries in these exploratory data.

### Search interest and total pickleball-related injuries

Annual total pickleball-related injury estimates correlated strongly with general pickleball Google Trends interest (R=0.96) and Pickleball Wikipedia pageviews (R=0.98). The sourdough control also correlated with the same annual injury estimates (R=0.76, **Figure 2A**). Monthly-level Pearson correlation coefficients across the 84 overlap months ranged from 0.34 to 0.59 for the total-injury attention signals (84 months, 2017-2023, when the injury, search, pageview and news series all overlap), indicating that the strength of the reported associations depended on temporal aggregation.

**Figure 2.**
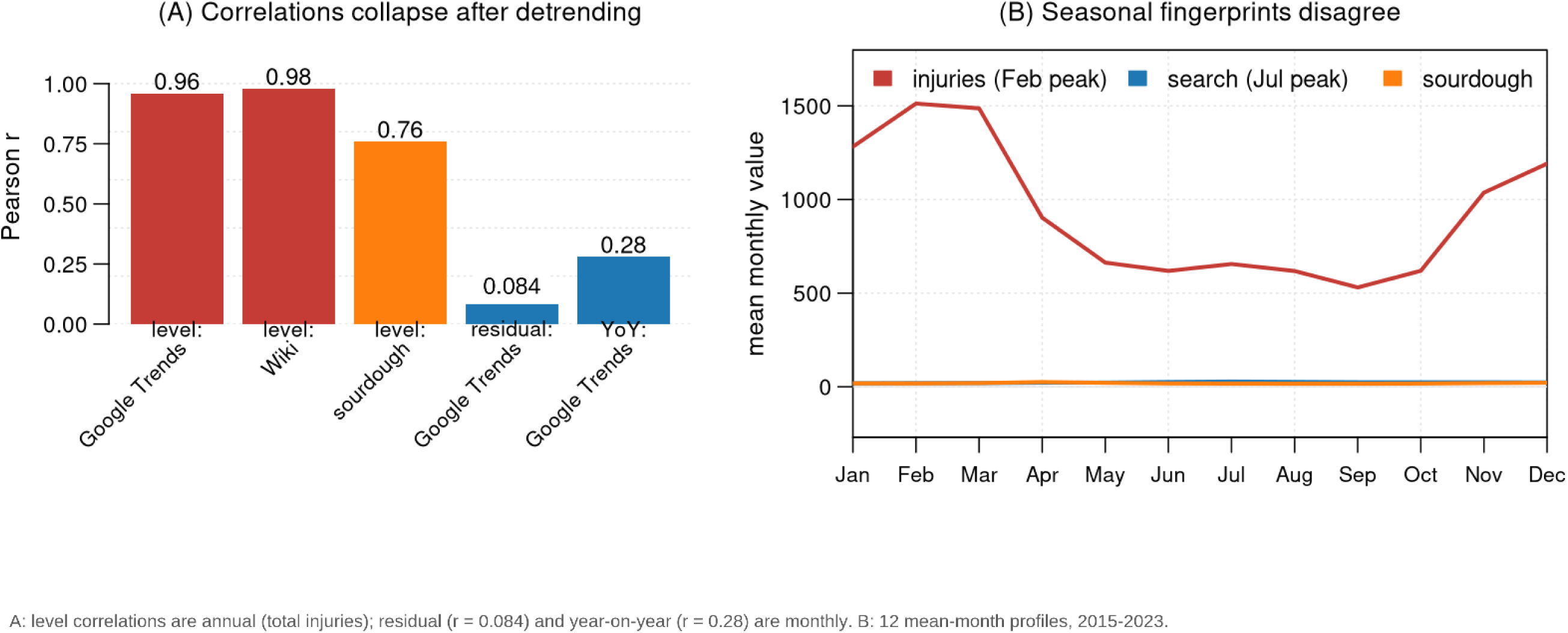
Pickleball injuries and internet attention before and after accounting for time trends. (A) Annual correlations of total pickleball-related injuries with general pickleball Google Trends interest, Pickleball Wikipedia pageviews, and the sourdough control, alongside monthly pickleball injury and Google Trends residual (R=0.084) and year-on-year (R=0.28) correlations. (B) Twelve-month mean profiles of raw monthly values, 2015 to 2023: injuries peak in February and searches in July. These raw profiles are not standardized STL components. Full seasonal-component and profile correlations are reported in the text with their estimands labelled.

After removing trend and seasonality, the monthly Google search-pickleball injury residual correlation was R=0.084. The year-on-year correlation was R=0.28 over 96 months (**Figure 2A**). Seasonal patterns also differed where injuries peaked in February while pickleball searches peaked in July. On the full 108-month seasonal components, the seasonal correlation with injuries was -0.35 for pickleball searches and +0.81 for sourdough searches; on 12-month mean profiles the corresponding values were -0.40 and +0.96 (**Figure 2B** shows the raw mean-month profiles). Sourdough baking itself surged during the 2020 lockdowns, so the placebo shares the period effect.

Pickleball news coverage rose about five-fold during the study window (summed GDELT daily volume-intensity shares, 1.75 in 2017 to 9.27 in 2023) and correlated with total pickleball-related injuries in annual levels (R=0.92). Google search interest for "pickleball" and pickleball news coverage tracked each other at every level (R=0.85 in monthly levels and R=0.64 in year-on-year changes). However, year-on-year correlations with total injuries were smaller, at 0.25 for searches and 0.20 for news (**Figure 3A** and **3B**). In these comparisons, search interest tracked news coverage more closely than it tracked injuries.

**Figure 3.**
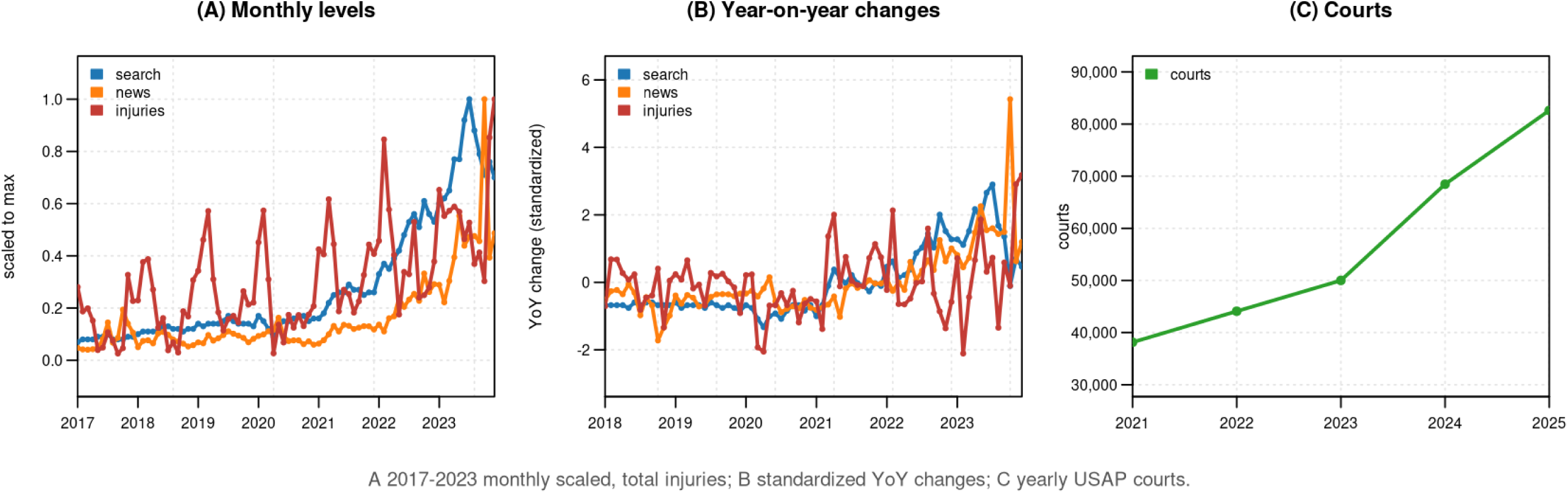
Search interest, news coverage, and contextual growth measures. (A) Monthly general pickleball Google search interest, pickleball GDELT news coverage, and total pickleball-related injuries during 2017-2023 (news coverage begins in 2017). (B) Year-on-year changes (standardized for display; correlations unchanged): search-news R=0.64, injury-search R=0.25, and injury-news R=0.20. (C) USA Pickleball court totals rise from 38,140 in 2021 to 82,613 in 2025. This contextual series extends beyond the injury-study period. Source and retrieval details are described in the Methods.

The contextual series also increased as the USA Pickleball court totals rose from 38,140 in 2021 to 82,613 in 2025 (**Figure 3C**). These observations describe growth beyond the injury-study period and were not used to estimate injury risk.

### Sensitivity and contextual checks

Repeating the monthly analyses using unweighted total case counts gave an injury-search residual correlation of 0.16 and a year-on-year correlation of 0.38 on the archived search draw. On the full seasonal components, unweighted seasonal correlations were -0.27 for pickleball searches and 0.79 for sourdough searches, with injuries peaking in February. The previously reported 0.17 year-on-year value was the orthopedic-only series and is not used here as a total-case result. As a robustness check, Spearman rank correlations were in the same direction (year-on-year injury–search ρ=0.35, nominal P<0.001 without multiplicity correction; monthly levels ρ=0.68). Thus, the associations were attenuated relative to annual levels in both weighted and unweighted analyses, rather than disappearing.

In 2020, consumer-product NEISS sample records fell by 13.8 percent and adjudicated pickleball cases by 15.7 percent (**Figure 1A**). Pickleball’s share of sample records was therefore similar to that in the previous year (ratio 0.98). These changes show that the pickleball decline occurred alongside a broader decline in recorded emergency department attendance. The dip is consistent with both reduced play during lockdowns and reduced emergency-department attendance; these data cannot separate reduced exposure, fewer injuries, and changes in care seeking.

## DISCUSSION

In this work, we asked whether the pickleball boom has raised injury risk for adults, and in particular, adults aged 60 to 79 years. The available data do not suggest that this is the case, as denominator-matched analyses did not detect a statistically significant increase in risk. However, the variability in the data cannot definitively rule out an increase in individual injury risk. Separately, surface-level strong annual associations between total pickleball-related injuries and general pickleball internet attention were weak after accounting for trend and seasonality.

Pickleball-related injury counts describe the burden on patients and emergency departments. They cannot, without an appropriate exposure denominator, show that the sport has become more hazardous. We use a single total-player denominator, since the public snapshots we located do not provide a harmonized annual frequency-specific series, matched 60 to 79 denominators, or person-hours with uncertainty. As a result, non-significant trends should not be read as evidence of safety or of stable risk.

The search analysis illustrates a different problem: a strong association in levels can offer little information about changes in the outcome. A correlation near 0.9 in levels is the product of two lines that both rise, and was substantially attenuated after the trend and seasonality they share were removed. A sourdough negative control fits the injury season better than pickleball does. News coverage, a second measure of public attention, stays coupled to search interest in year-on-year changes while neither stays coupled to injuries, which suggests that the search signal is a measure of attention, rather than a count of injuries. This recalls Google Flu Trends: a search index is a sampled, rescaled, and media-sensitive quantity, and, in this study, a poor thermometer for pickleball injury counts <u>[25–27]</u>.

Our work identified two distinct errors in extrapolating from the surge in pickleball interest to increased injuries. The first is conceptual. A count is not a risk, and no analysis can compensate for a missing denominator. The second is statistical. Two series that climb together will correlate regardless of any causal link (i.e., pickleball searches and injuries). The safeguards are straightforward. Remove the shared trend, test against a negative control (i.e., sourdough), and divide by the number of people at risk before drawing conclusions.

### Comparison with other studies

Our rising injury estimates are consistent with the NEISS literature, showing pickleball emergency visits rising steeply with a skew toward adults aged 60 years or older <u>[2–7]</u>. This national NEISS-based pickleball-injury study uses SFIA participation denominators rather than raw or census-based counts (Owoeye and colleagues used 1,758 surveyed players as an injury-prevalence denominator in a 2025 survey <u>[31]</u>); this addresses a question that raw counts or census population denominators cannot settle. One earlier study separately reported that search volume and injury counts rose together <u>[10]</u>. However, when we applied standard time series checks to the same kind of comparison, the association was small (year-on-year R=0.28).

Howe and colleagues’ Pokemon GO study <u>[28]</u> is a different design from ours: an initial increase in daily steps after installation waned over six weeks in a difference-in-differences analysis. We cite it only as an example of an early association attenuating over time, not as a detrending analysis. In our study, the search correlation followed the same arc: from 0.96 to 0.98 in annual levels to small after removing trend and seasonality (monthly residual R=0.084; year-on-year R=0.28 over 96 months, 2015-2023).

### Strengths and limitations

Strengths of our work include case-level surveillance data, design-based confidence intervals, a single verified participation denominator, and comparisons across several attention signals and a negative control <u>[13]</u>. Repeating monthly analysis with unweighted counts provided an additional check on sensitivity to the weighting used to extrapolate NEISS to the general US population.

Precision is the principal limitation of the rate analysis. Seven annual observations and wide injury-estimate confidence intervals limit inference about changes in risk. The analysis does not quantify how large a change can be excluded. SFIA participation estimates are survey-based totals without published uncertainty, and the survey methods may have changed over the window <u>[1]</u>; our rate confidence interval propagates only the injury-estimate uncertainty and therefore understates total uncertainty. Core-player splits and modeled person-hours were dropped for the same reason; they remain future work with better data.

The monthly analyses are exploratory because 87 of 120 months contained fewer than 20 cases. Narrative-based case identification can misclassify injuries; the diagnosis-code mapping was confirmed against the January 2023 CPSC manual, but narrative misclassification remains possible. Google Trends is sampled and rescaled, and the sparse court series provides context rather than injury validation. Pandemic changes in recorded attendance cannot separate changes in exposure, injury occurrence, and care seeking. Finally, these ecological comparisons cannot establish individual-level associations or causal effects <u>[29]</u>. Age-stratified and body-part-specific rates would require compatible age-specific participation denominators, which is left for future work.

### Conclusions

In summary, we do not find compelling evidence of a pickleball-induced injury epidemic. While the rising injury burden deserves attention <u>[30]</u>, when normalized to the number of participants, we did not detect a temporal trend. Rising search interest did not reliably track changes in total pickleball-related injuries in these analyses. Hence, before interpreting a rising trend, examine its denominator, remove shared trends, and test findings against a negative control.

## DATA AVAILABILITY

All data was conducted on publicly available data, as detailed in the Methods. The code and aggregate results are available from the Corresponding author on reasonable request.

## USE OF ARTIFICIAL INTELLIGENCE

Muse Wonder was used for analyses, drafting of tables and figures, and drafting of text. The authors reviewed all work and take responsibility for the validity of analyses.

## Supplementary material

**Supplementary Table S1.** STROBE checklist adapted to ecological time-series.

| # | STROBE item | Location in manuscript |
| --- | --- | --- |
| 1a | Design in title/abstract | Title; Abstract DESIGN |
| 1b | Informative balanced abstract | Abstract incl. stability caveat |
| 2 | Background/rationale | Introduction |
| 3 | Objectives | Objectives paragraph |
| 4 | Study design | Analysis paragraph |
| 5 | Setting | US, 2014–2023; retrieval windows |
| 6a | Participants (eligibility) | Validated case definition; Table S2; older adults 60+ |
| 7 | Variables | Outcomes; SFIA totals; signals; sourdough control |
| 8 | Data sources/measurement | Methods; windows + receipts; SFIA provenance |
| 9 | Bias | Limitations |
| 10 | Study size | 1,738 cases (921 orthopedic); series lengths |
| 11 | Quantitative variables | Age bands; STL/YoY constructions |
| 12 | Statistical methods | OLS t-inference; Pearson + Spearman; STL params; ADF |
| 13–15 | Participants/descriptives/outcomes | Results; Table 1; Tables S3 and S4; Figures 1 to 3, S1 and S2 |
| 16 | Main results | Rate null; detrended search null (adjusted + unadjusted) |
| 17 | Other analyses | Unweighted; ortho-only; sex/age/half/subtype splits |
| 18 | Key results | Discussion opening |
| 19 | Limitations | Limitations section |
| 20 | Interpretation | Discussion; Conclusions |
| 21 | Generalisability | US ED-treated injuries; scope limits stated |
| 22 | Funding | Sources of Funding (Meta Platforms, Inc.) |

**Supplementary Table S2.** NEISS narrative string review. Naive case-insensitive screen for PICKLE over NEISS Narrative 1 + 2 (2014–2023) returned 1,958 records; review found 11% false positives (pickle jars / food preparation). The rule retained 1,739 of 1,958 records (yield 88.8%; no independent gold-standard adjudication was performed, so this yield is not a validated positive predictive value). Examples: *injured wrist opening pickle jar* → exclude; *cut finger slicing a pickle* → exclude; *playing pickleball, fell on court* → keep. Term lists verbatim from the replication extractor (neiss_extract.py in the recovered replication package).

| # | Screen / rule | Strings (case-insensitive) | Decision | n |
| --- | --- | --- | --- | --- |
| 0 | Naive screen | PICKLE over Narrative 1 + 2 | Review set | 1,958 |
| 1 | Explicit game token, no food term | pickleball, pickle ball, picklebal, pickle-ball | Keep | ⊆ 1,739 |
| 2 | Pickle + playing term, no food term | court, paddle, serve, volley, dink, kitchen, rally, doubles, singles, net, match, tournament | Keep | ⊆ 1,739 |
| 3 | Food/jar evidence | jar, food, slic*, eating, preparat*, preserv*, cucumber, dill, brine, canning, gherkin, relish | Exclude | ≈ 219 (11%) |
| 4 | Food term + game token | — | Manual review, not auto-kept | 0 by rule |
| 5 | pickle with no other context | — | Manual review, not auto-kept | 0 by rule |
| 6 | Validated set (all keeps) | — | Keep | 1,739 |
| 7 | Analyzed set (after adjudication) | case 140424871 removed | Analyze | 1,738 |

**Supplementary Table S3.** Orthopaedic pickleball injuries, NEISS 2014–2023. Annual orthopaedic subset (fracture, dislocation, sprain/strain; 921 of 1,738 adjudicated cases) with design-based (Taylor linearization) 95 percent confidence intervals and CPSC stability flags.

| Year | Sample cases (N) | National estimate | 95% CI | CV (%) | CPSC stability |
| --- | --- | --- | --- | --- | --- |
| 2014 | 15 | 870 | 136 to 1,605 | 43.1 | Unstable |
| 2015 | 32 | 2,231 | 143 to 4,318 | 47.7 | Unstable |
| 2016 | 40 | 2,949 | 490 to 5,407 | 42.5 | Unstable |
| 2017 | 59 | 3,668 | 0 to 7,449 | 52.6 | Unstable |
| 2018 | 77 | 4,953 | 0 to 10,002 | 52.0 | Unstable |
| 2019 | 92 | 6,138 | 0 to 13,095 | 57.8 | Unstable |
| 2020 | 92 | 5,618 | 0 to 11,759 | 55.8 | Unstable |
| 2021 | 162 | 8,509 | 0 to 17,619 | 54.6 | Unstable |
| 2022 | 140 | 8,479 | 1,657 to 15,300 | 41.0 | Unstable |
| 2023 | 212 | 10,290 | 5,488 to 15,092 | 23.8 | Stable |

**Supplementary Table S4.** NEISS code mapping as used in this study. Diagnosis codes 57/55/64 → fracture / dislocation / sprain-strain; leading fracture body-part codes 34 → wrist, 79 → lower trunk, 33 → lower arm, 37 → ankle. Confirmed against the CPSC NEISS Coding Manual (January 2023). An independent narrative-text audit supports the mapping: 100% of code-57 cases carry fracture tokens, 83% of code-64 sprain/strain tokens, 91% of code-55 dislocation tokens; wrist mentions dominate code 34 (128 of 144 fracture cases, 89%), all 95 cases carrying body-part code 37 across all diagnoses mentioned ankle (code 37 accounts for 3.2% of the 472 fractures in the table below), while 23 additional ankle-mention cases had another primary body-part code.

| Code type | Code | Meaning as used |
| --- | --- | --- |
| Diagnosis | 57 | Fracture |
| Diagnosis | 55 | Dislocation |
| Diagnosis | 64 | Sprain / strain |
| Body part | 34 | Wrist (30.5% of fractures) |
| Body part | 79 | Lower trunk (15.7%) |
| Body part | 33 | Lower arm (14.4%) |
| Body part | 37 | Ankle (3.2%) |

**Supplementary Table S5.** Injury-to-participation measures with 95 percent confidence intervals, 2017–2023. Rates are per 1,000 players. Rates for 2014–2016 are not computed because no participation denominators were verified and included for those years in this analysis. Lower bounds below zero are shown as —. The denominator is treated as fixed, so intervals are lower bounds on uncertainty. Source: supplement_rate_cis.csv in the replication package.

| Year | Total-player rate (95% CI) |
| --- | --- |
| 2017 | 1.96 (0.03 to 3.89) |
| 2018 | 2.52 (— to 5.05) |
| 2019 | 3.19 (— to 6.52) |
| 2020 | 2.15 (— to 4.36) |
| 2021 | 3.09 (— to 6.30) |
| 2022 | 1.91 (0.34 to 3.48) |
| 2023 | 1.79 (0.77 to 2.80) |

**Supplementary Figure S1.**
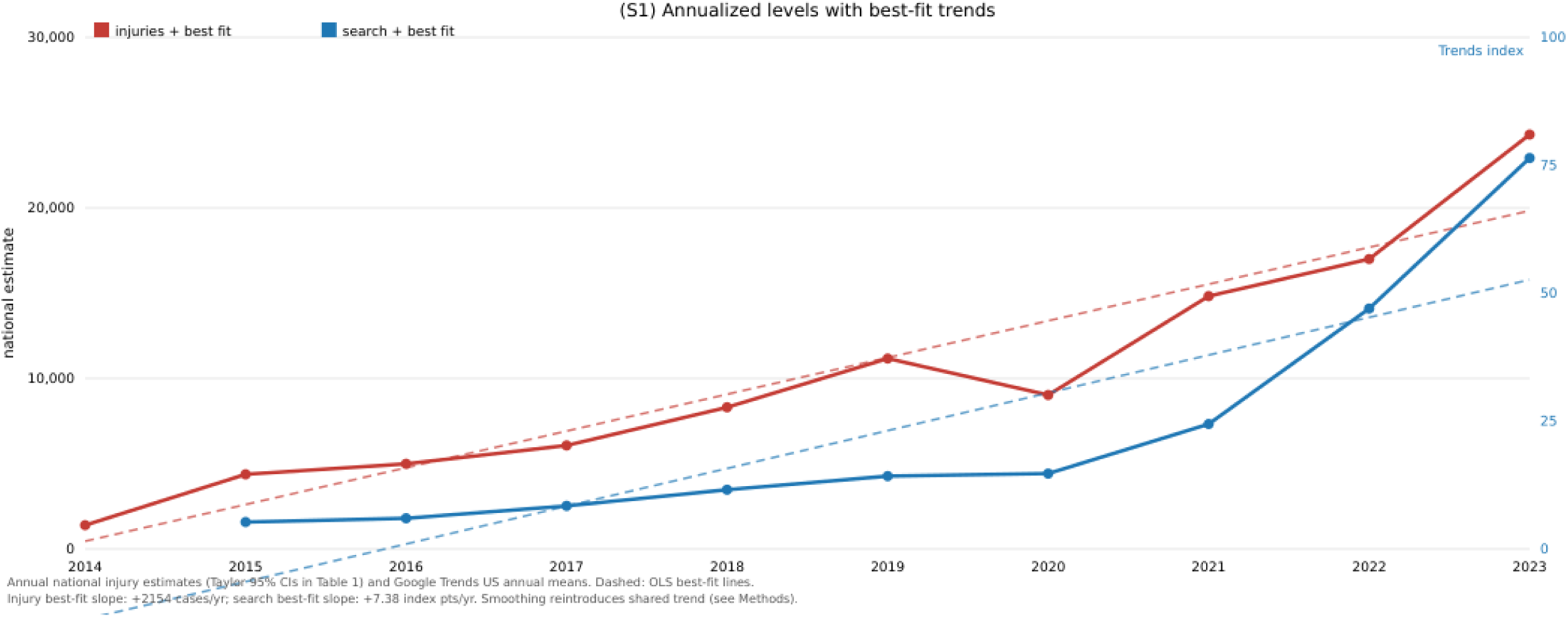
Annualized levels with OLS best-fit trends (injury slope +2,154 cases/year; search +7.38 index points/year). Dashed lines are OLS fits; smoothing reintroduces the shared trend whose removal is the point of the check (see Methods).

**Supplementary Figure S2.**
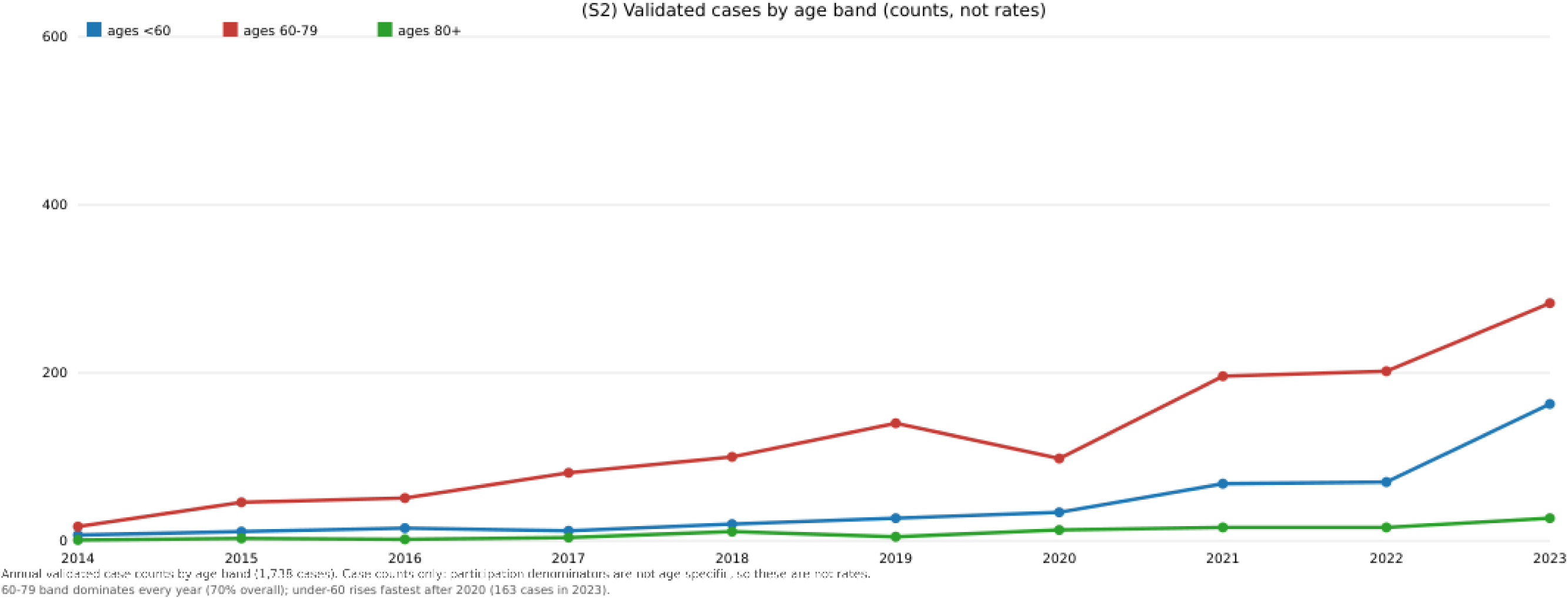
Annual adjudicated case counts by age band (1,738 cases). Counts only: participation denominators are not age-specific, so these are not rates. The 60–79 band dominates every year (70% overall); under-60 cases rise fastest after 2020 (163 in 2023).

